# Characteristics of SARS-CoV-2 Positive Individuals in California From Two Periods During Notable Decline in Incident Infection

**DOI:** 10.1101/2021.05.11.21257008

**Authors:** Lao-Tzu Allan-Blitz, Isaac Turner, Fred Hertlein, Jeffrey D. Klausner

## Abstract

Despite declining SARS-CoV-2 incidence, continued epidemic monitoring is warranted. We collected SARS-CoV-2 test results from 150 drive-through testing centers across California from two observation periods: February 23^rd^-March 3^rd^ 2021 and April 15^th^-April 30^th^ 2021. We assessed SARS-CoV-2 positivity, stratified by Hispanic heritage among sociodemographic characteristics and potential exposures. We analyzed 114,789 test results (5.1% and 2.6% positive during the respective observation periods). Nearly half of all positive tests were among testers reporting a recent exposure (48.8% and 45.3% during the respective observation periods). Those findings may provide insight into evolving local transmission dynamics and support targeted public health strategies.

## Introduction

Over the last few months, the daily incidence of SARS-CoV-2 cases has declined sharply in California (1), as well as in many parts of the United States. The cause of the dramatic decline is likely multifactorial but greatly influenced by population-level immunity due to prior infection and increasing vaccination coverage (see Figure) (2).

**Figure:**
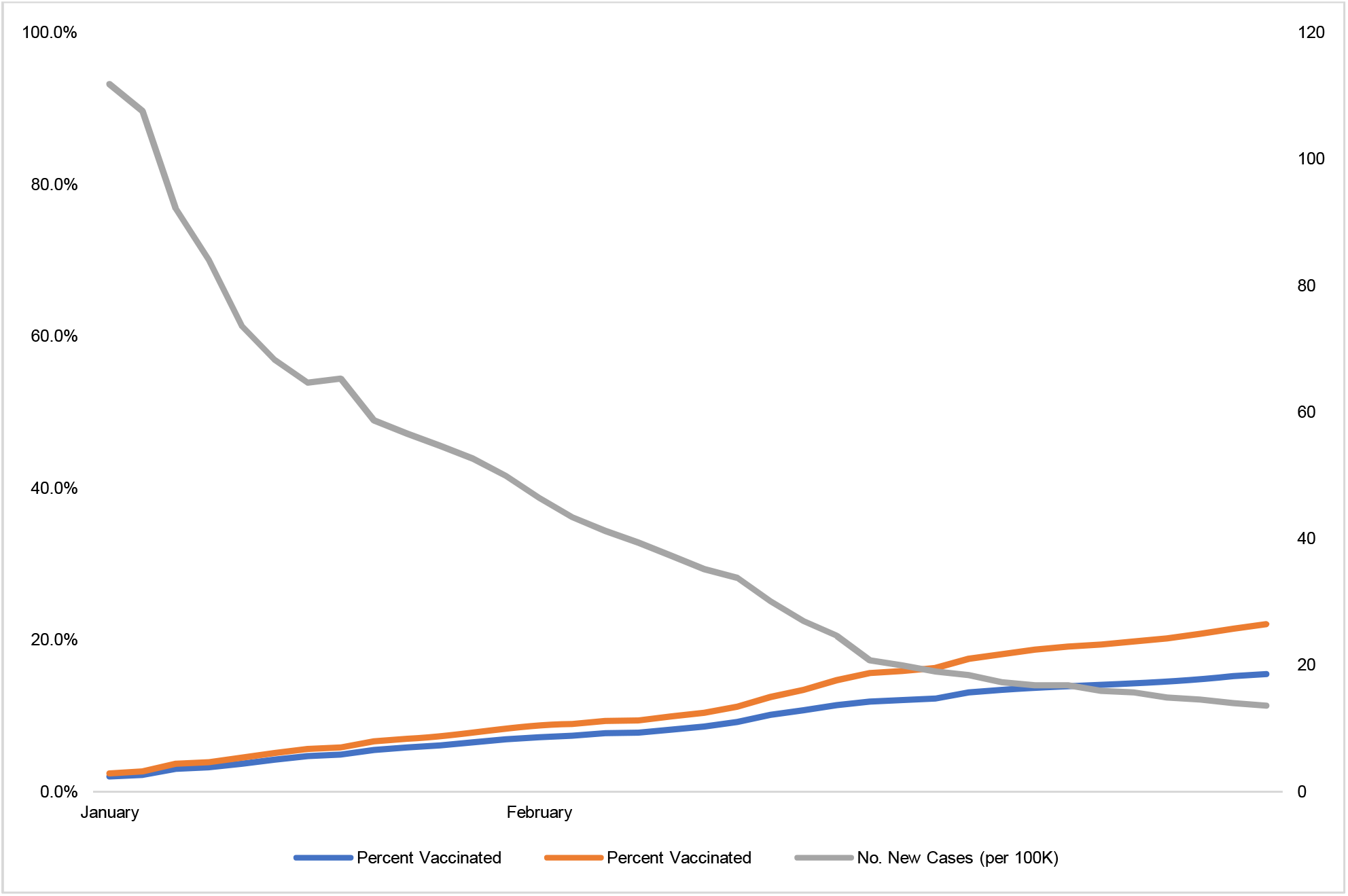
New SARS-CoV-2 Infections Against Percent Vaccinated in California, January - March 2021. This is a figure shows percent vaccinated (by first and second dose) and new SARS-CoV-2 cases per 100,000 individuals in California between January to March 2021. Data extrapolated from U.S. COVID Risk & Vaccine Tracker (2).

SARS-CoV-2, however, continues to be transmitted heterogeneously among different subsets of the population (3), and over the last few months several states reported resurging case counts (1). Fears that certain variants of SARS-CoV-2 (4) may escape immunity warrant continued epidemic monitoring. We aimed to characterize characteristics of those with SARS-CoV-2 positive test results during the recent decline in cases in California to understand current viral transmission.

## Methods

We evaluated SARS-CoV-2 polymerase chain reaction (PCR) test results from a large testing program in California at two time periods: 1) February 23^rd^ - March 3^rd^ 2021, and 2) between April 15^th^ - April 30^th^ 2021. Individuals presented for testing to any of 150 drive-through publicly accessible sites located in various counties and cities including Los Angeles, Riverside, San Mateo, Berkeley, Menlo Park, Maywood, and Rancho Mirage. Those who completed testing were asked via a confidential online survey if in the last 14 days they had been contacted by local public health authorities about a known SARS-CoV-2 exposure, they visited any of a list of public places, or spent time with five or more strangers, as well as demographic information, employment information, and report of any symptoms at the time of testing.

Self-collected oral swabs, which have been shown to perform similarly to clinician-collected nasopharyngeal swabs (5), were processed with standard PCR methods using a modified Center for Disease Control and Prevention testing protocol as has been previously reported (6). We then conducted a cross-sectional analysis to determine the frequency of infection among testers based on each of the above characteristics. Further, we calculated positivity ratios by various characteristics to evaluate trends in SARS-CoV-2 positivity during a period of notable decline in incident infection. To account for confounding of Hispanic heritage, given the reports of the disproportionate burden of infection among Hispanic persons (7), we stratified our analysis by heritage. The Mass General Brigham institutional review board deemed the analysis of de-identified data did not constitute human subjects’ research (2020P003530). All analyses were conducted using STATA 15.1 (StataCorp, College Station, TX).

## Results

We analyzed 114,789 test results (see Table). Of 52,9078 results between February 23^rd^ - March 3^rd^,2021 2,679 (5.1%) were positive. Of 61,882 test results from April 15^th^ - 30^th^, 1,579 (2.6%) were positive. Testers included 47.9% who identified with Hispanic heritage, and 54.6% reported female sex.

**Table:**
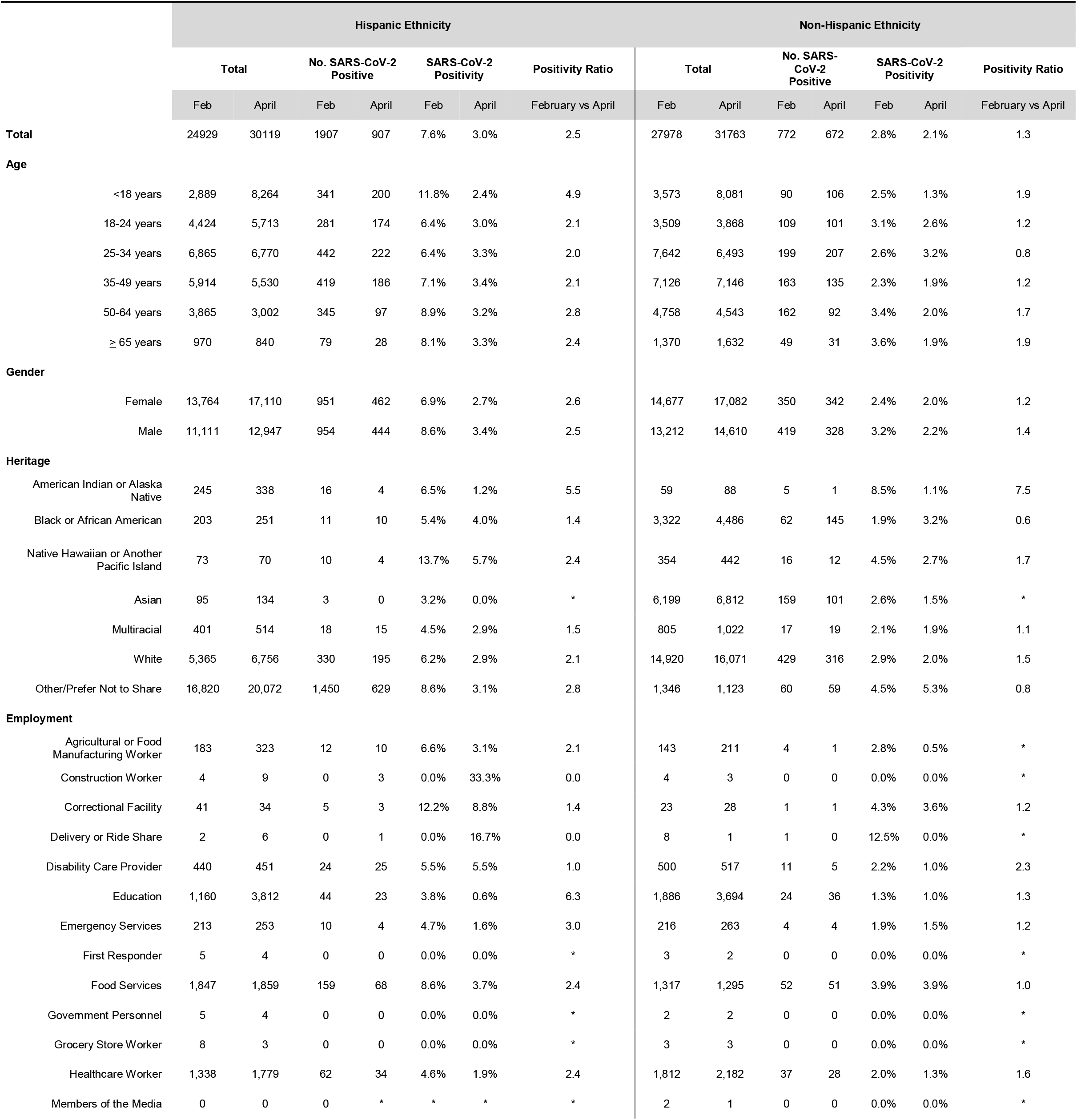
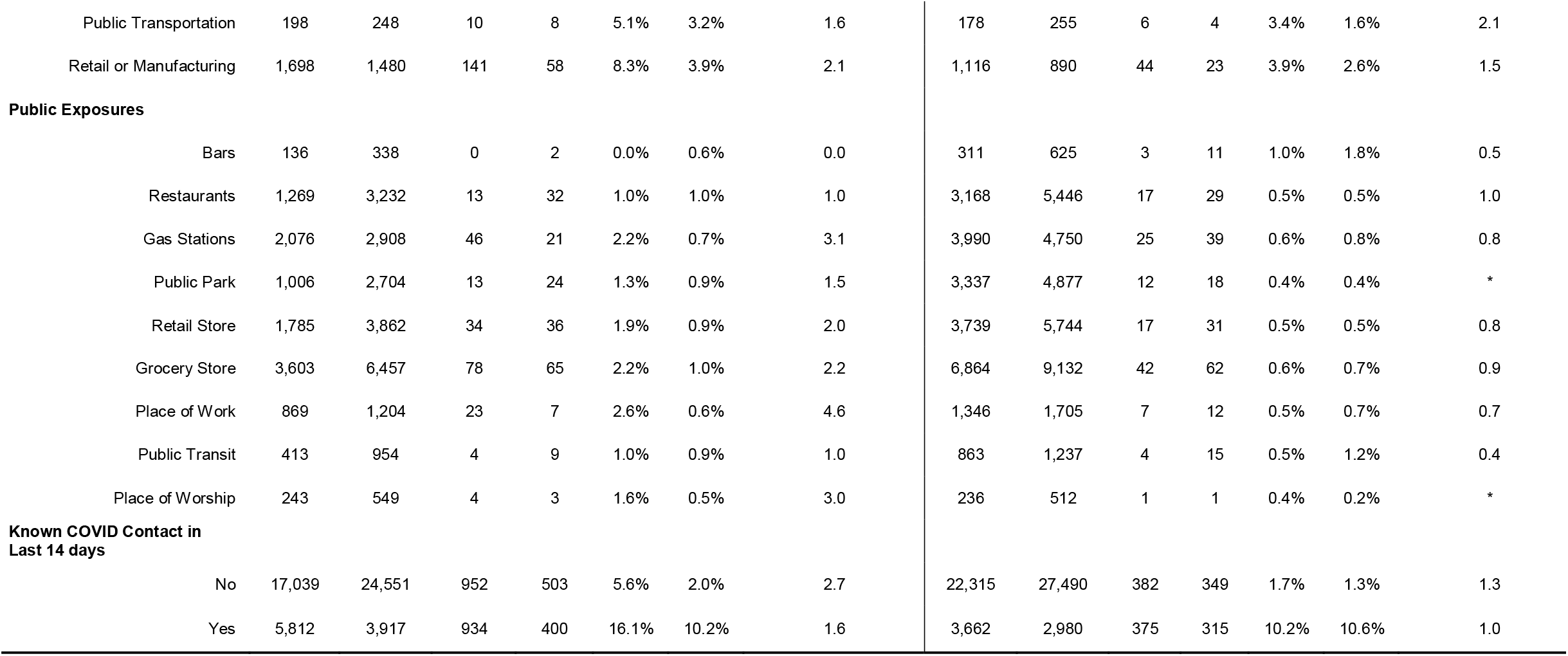
**SARS-CoV-2 Positivity Among Hispanics Individuals From California Comparing February 23rd - March 3rd and April 15th - April 30th, 2021**

|  | Hispanic Ethnicity |  |  |  |  |  |  |  | Non-Hispanic Ethnicity |  |  |  |  |  |  |
| --- | --- | --- | --- | --- | --- | --- | --- | --- | --- | --- | --- | --- | --- | --- | --- |
|  | Total |  | No. SARS-CoV-2 Positive |  | SARS-CoV-2 Positivity |  | Positivity Ratio |  | Total |  | No. SARS-CoV-2 Positive |  | SARS-CoV-2 Positivity |  | Positivity Ratio |
|  | Feb | April | Feb | April | Feb | April | February vs April |  | Feb | April | Feb | April | Feb | April | February vs April |
| **Total** | 24929 | 30119 | 1907 | 907 | 7.6% | 3.0% | 2.5 |  | 27978 | 31763 | 772 | 672 | 2.8% | 2.1% | 1.3 |
| **Age** |  |  |  |  |  |  |  |  |  |  |  |  |  |  |  |
| <18 years | 2,889 | 8,264 | 341 | 200 | 11.8% | 2.4% | 4.9 |  | 3,573 | 8,081 | 90 | 106 | 2.5% | 1.3% | 1.9 |
| 18-24 years | 4,424 | 5,713 | 281 | 174 | 6.4% | 3.0% | 2.1 |  | 3,509 | 3,868 | 109 | 101 | 3.1% | 2.6% | 1.2 |
| 25-34 years | 6,865 | 6,770 | 442 | 222 | 6.4% | 3.3% | 2.0 |  | 7,642 | 6,493 | 199 | 207 | 2.6% | 3.2% | 0.8 |
| 35-49 years | 5,914 | 5,530 | 419 | 186 | 7.1% | 3.4% | 2.1 |  | 7,126 | 7,146 | 163 | 135 | 2.3% | 1.9% | 1.2 |
| 50-64 years | 3,865 | 3,002 | 345 | 97 | 8.9% | 3.2% | 2.8 |  | 4,758 | 4,543 | 162 | 92 | 3.4% | 2.0% | 1.7 |
| ≥ 65 years | 970 | 840 | 79 | 28 | 8.1% | 3.3% | 2.4 |  | 1,370 | 1,632 | 49 | 31 | 3.6% | 1.9% | 1.9 |
| **Gender** |  |  |  |  |  |  |  |  |  |  |  |  |  |  |  |
| Female | 13,764 | 17,110 | 951 | 462 | 6.9% | 2.7% | 2.6 |  | 14,677 | 17,082 | 350 | 342 | 2.4% | 2.0% | 1.2 |
| Male | 11,111 | 12,947 | 954 | 444 | 8.6% | 3.4% | 2.5 |  | 13,212 | 14,610 | 419 | 328 | 3.2% | 2.2% | 1.4 |
| **Heritage** |  |  |  |  |  |  |  |  |  |  |  |  |  |  |  |
| American Indian or Alaska Native | 245 | 338 | 16 | 4 | 6.5% | 1.2% | 5.5 |  | 59 | 88 | 5 | 1 | 8.5% | 1.1% | 7.5 |
| Black or African American | 203 | 251 | 11 | 10 | 5.4% | 4.0% | 1.4 |  | 3,322 | 4,486 | 62 | 145 | 1.9% | 3.2% | 0.6 |
| Native Hawaiian or Another Pacific Island | 73 | 70 | 10 | 4 | 13.7% | 5.7% | 2.4 |  | 354 | 442 | 16 | 12 | 4.5% | 2.7% | 1.7 |
| Asian | 95 | 134 | 3 | 0 | 3.2% | 0.0% | * |  | 6,199 | 6,812 | 159 | 101 | 2.6% | 1.5% | * |
| Multiracial | 401 | 514 | 18 | 15 | 4.5% | 2.9% | 1.5 |  | 805 | 1,022 | 17 | 19 | 2.1% | 1.9% | 1.1 |
| White | 5,365 | 6,756 | 330 | 195 | 6.2% | 2.9% | 2.1 |  | 14,920 | 16,071 | 429 | 316 | 2.9% | 2.0% | 1.5 |
| Other/Prefer Not to Share | 16,820 | 20,072 | 1,450 | 629 | 8.6% | 3.1% | 2.8 |  | 1,346 | 1,123 | 60 | 59 | 4.5% | 5.3% | 0.8 |
| **Employment** |  |  |  |  |  |  |  |  |  |  |  |  |  |  |  |
| Agricultural or Food Manufacturing Worker | 183 | 323 | 12 | 10 | 6.6% | 3.1% | 2.1 |  | 143 | 211 | 4 | 1 | 2.8% | 0.5% | * |
| Construction Worker | 4 | 9 | 0 | 3 | 0.0% | 33.3% | 0.0 |  | 4 | 3 | 0 | 0 | 0.0% | 0.0% | * |
| Correctional Facility | 41 | 34 | 5 | 3 | 12.2% | 8.8% | 1.4 |  | 23 | 28 | 1 | 1 | 4.3% | 3.6% | 1.2 |
| Delivery or Ride Share | 2 | 6 | 0 | 1 | 0.0% | 16.7% | 0.0 |  | 8 | 1 | 1 | 0 | 12.5% | 0.0% | * |
| Disability Care Provider | 440 | 451 | 24 | 25 | 5.5% | 5.5% | 1.0 |  | 500 | 517 | 11 | 5 | 2.2% | 1.0% | 2.3 |
| Education | 1,160 | 3,812 | 44 | 23 | 3.8% | 0.6% | 6.3 |  | 1,886 | 3,694 | 24 | 36 | 1.3% | 1.0% | 1.3 |
| Emergency Services | 213 | 253 | 10 | 4 | 4.7% | 1.6% | 3.0 |  | 216 | 263 | 4 | 4 | 1.9% | 1.5% | 1.2 |
| First Responder | 5 | 4 | 0 | 0 | 0.0% | 0.0% | * |  | 3 | 2 | 0 | 0 | 0.0% | 0.0% | * |
| Food Services | 1,847 | 1,859 | 159 | 68 | 8.6% | 3.7% | 2.4 |  | 1,317 | 1,295 | 52 | 51 | 3.9% | 3.9% | 1.0 |
| Government Personnel | 5 | 4 | 0 | 0 | 0.0% | 0.0% | * |  | 2 | 2 | 0 | 0 | 0.0% | 0.0% | * |
| Grocery Store Worker | 8 | 3 | 0 | 0 | 0.0% | 0.0% | * |  | 3 | 3 | 0 | 0 | 0.0% | 0.0% | * |
| Healthcare Worker | 1,338 | 1,779 | 62 | 34 | 4.6% | 1.9% | 2.4 |  | 1,812 | 2,182 | 37 | 28 | 2.0% | 1.3% | 1.6 |
| Members of the Media | 0 | 0 | 0 | * | * | * | * |  | 2 | 1 | 0 | 0 | 0.0% | 0.0% | * |

|  |  |  |  |  |  |  |  |  |  |  |  |  |  |  |
| --- | --- | --- | --- | --- | --- | --- | --- | --- | --- | --- | --- | --- | --- | --- |
| Public Transportation | 198 | 248 | 10 | 8 | 5.1% | 3.2% | 1.6 | 178 | 255 | 6 | 4 | 3.4% | 1.6% | 2.1 |
| Retail or Manufacturing | 1,698 | 1,480 | 141 | 58 | 8.3% | 3.9% | 2.1 | 1,116 | 890 | 44 | 23 | 3.9% | 2.6% | 1.5 |
| **Public Exposures** |  |  |  |  |  |  |  |  |  |  |  |  |  |  |
| Bars | 136 | 338 | 0 | 2 | 0.0% | 0.6% | 0.0 | 311 | 625 | 3 | 11 | 1.0% | 1.8% | 0.5 |
| Restaurants | 1,269 | 3,232 | 13 | 32 | 1.0% | 1.0% | 1.0 | 3,168 | 5,446 | 17 | 29 | 0.5% | 0.5% | 1.0 |
| Gas Stations | 2,076 | 2,908 | 46 | 21 | 2.2% | 0.7% | 3.1 | 3,990 | 4,750 | 25 | 39 | 0.6% | 0.8% | 0.8 |
| Public Park | 1,006 | 2,704 | 13 | 24 | 1.3% | 0.9% | 1.5 | 3,337 | 4,877 | 12 | 18 | 0.4% | 0.4% | * |
| Retail Store | 1,785 | 3,862 | 34 | 36 | 1.9% | 0.9% | 2.0 | 3,739 | 5,744 | 17 | 31 | 0.5% | 0.5% | 0.8 |
| Grocery Store | 3,603 | 6,457 | 78 | 65 | 2.2% | 1.0% | 2.2 | 6,864 | 9,132 | 42 | 62 | 0.6% | 0.7% | 0.9 |
| Place of Work | 869 | 1,204 | 23 | 7 | 2.6% | 0.6% | 4.6 | 1,346 | 1,705 | 7 | 12 | 0.5% | 0.7% | 0.7 |
| Public Transit | 413 | 954 | 4 | 9 | 1.0% | 0.9% | 1.0 | 863 | 1,237 | 4 | 15 | 0.5% | 1.2% | 0.4 |
| Place of Worship | 243 | 549 | 4 | 3 | 1.6% | 0.5% | 3.0 | 236 | 512 | 1 | 1 | 0.4% | 0.2% | * |
| **Known COVID Contact in Last 14 days** |  |  |  |  |  |  |  |  |  |  |  |  |  |  |
| No | 17,039 | 24,551 | 952 | 503 | 5.6% | 2.0% | 2.7 | 22,315 | 27,490 | 382 | 349 | 1.7% | 1.3% | 1.3 |
| Yes | 5,812 | 3,917 | 934 | 400 | 16.1% | 10.2% | 1.6 | 3,662 | 2,980 | 375 | 315 | 10.2% | 10.6% | 1.0 |

In the first period, the positivity of SARS-CoV-2 infection was higher among Hispanic (7.6%) compared to non-Hispanic (2.8%) testers (*p-*value<0.001). In the second period, the positivity of SARS-CoV-2 infection among Hispanic testers was 3.0% compared to 2.0% among non-Hispanic Individuals (*p-*value=0.09). Of all who tested positive in the first period 1,309 (48.8%) reported contact with an individual known to be infected with SARS-CoV-2 in the last 14 days. In the second period, a similar proportion, 715 (45.3%) of the 1,579 individuals who tested positive reported exposure to a known contact in the last 14 days.

Among Hispanic testers during the first period, we found a high positivity of infection among children (11.8%) and those who reported mixed Native Hawaiian or Pacific Island and Hispanic heritage (6.5%). We found consistently elevated positivity through both periods among individuals reporting any known exposure in the past 14 days (16.1% and 10.2%, respectively), individuals reporting employment as disability care providers (5.5% and 5.5%, respectively), food service providers (8.6% and 3.7%, respectively), and employment in retail or manufacturing (8.3% and 3.9%, respectively).

Among non-Hispanic testers, we found consistently elevated SARS-CoV-2 positivity among individuals reporting employment in retail or manufacturing in both periods, periods (3.9% and 2.6%, respectively) as well as among testers reporting any known exposure in the past 14 days (10.2% and 10.6%, respectively).

## Discussion

We evaluated sociodemographic characteristics and potential exposures among those presenting for SARS-CoV-2 testing during two periods of notable decline in incident cases in California. The current trends in SARS-CoV-2 case rates across the United States are at an uncertain point, with several states reporting continued declines, while some are experiencing a resurgence (1). An epidemiologic understanding of those most at risk for infection is essential as the transmission dynamics shift, in order to guide future prevention efforts toward those at ongoing risk. Prevention programs should include vaccination, and among those unvaccinated continued employment-based regular testing, ventilation, indoor mask use and distancing. We identified notable SARS-CoV-2 positivity among testers of Hispanic heritage and those reporting a recent known exposure.

Nearly half of infections in our testing population were among individuals who reported contact with someone known to be infected with SARS-CoV-2 in the last 14 days. A finding which persisted through both periods. Thus, contact tracing efforts, perhaps now more than ever, are essential for identifying and isolating remaining cases, and testing and quarantine of those exposed(8). Incorporation of detailed exposure reporting into contact tracing efforts may also facilitate real-time monitoring of the variations in risk exposures. The collection of more specific exposure data from both infected and uninfected persons undergoing testing should be added as a crucial strategy moving forward (9). Additionally, continuing to encourage testing and vaccination among individuals with a known exposure will be essential.

We found high test positivity among Hispanic testers, and among children specifically during the first period, supporting the key role of within household transmission (10). Because schools were mostly closed during the initial observation period in California, and the reopening of schools predominantly overlapped with the second observation period, it is unlikely that school attendance is contributing to the continued spread of infection. Household crowding among Hispanic individuals is common with higher average number of individuals per room with multiple generations (11), likely contributing more substantially substantial intra-familial transmission.

The disparities noted in SARS-CoV-2 positivity among testers identifying as individuals of mixed minority heritage are consistent with recent studies (7, 12, 13), which likely reflects systemic structural inequities that continue to put certain populations at increased risk of exposure due to inadequate protections. Particularly among individuals of Hispanic heritage, types of employment likely play a key role in sustained transmission (14). Our findings suggest that prevention strategies may benefit from focusing on employers in particular those employing food service workers and disability care providers. Disability care providers are a particularly important population as the morbidity and mortality of SARS-CoV-2 infection are substantial among individuals of long-term care facilities (15). Thus, we encourage requiring vaccination among such employment categories. Further research is still needed to both clarify the specific exposures within different sub-populations that may contribute to sustained transmission, as well as to address the underlying structural inequities that result in the disparities identified.

Our study had several limitations. First, we analyzed laboratory-based testing data and could not account for individuals with repeat testing. Second, we were unable to collect detailed socioeconomic data like household size in order to control for confounding factors. Data collection was also incomplete for some fields making further statistical analyses and modeling not possible. However, the strengths of our study were the very large sample size, thus improving precision of our results, the unbiased collection of exposure data prior to receiving testing results, and the inclusion of numerous testing sites across California, improving the generalizability of our results.

## Conclusions

We report SARS-CoV-2 positivity by sociodemographic employment and various exposure characteristics in California from two observation periods during a notable decline in incident infection. We found higher SARS-CoV-2 positivity among Hispanic testers as well as testers with a known recent exposure, as well as variations in positivity by type of employment. Those findings are important because of the evolving epidemiology of those at risk as the incidence of infection decreases, thus providing the groundwork for future research and potentially informing public health strategies.

## Data Availability

The data is available upon request

## Funding and Acknowledgements

The authors would like to acknowledge the cities of Los Angeles, Riverside, San Mateo, Berkeley, Menlo Park, Maywood, and Rancho Mirage. There was no funding for this project.

## References

1. Centers for Disease Control and Prevention. COVID Data Tracker: United States COVID-19 Cases and Deaths by State. Last updated April 2nd 2021. Available at: https://covid.cdc.gov/covid-data-tracker/#cases_casesper100klast7days Accessed April 4th, 2021.

2. U.S. COVID Risk & Vaccine Tracker. Available at: https://covidactnow.org/?s=1793712 Last Accessed April 29th, 2021.

3. Centers for Disease Control and Prevention. COVID Data Tracker: Integrated County View. Available at: covid.cdc.gov/covid-data-tracker/?CDC_AA_refVal=https%3AY2F%2Fwww.cdc.gov%2Fcoronavirus%2F2019-ncov%2Fcases-updates%2Fcases-in-us.html#county-view Accessed 5/9/21. 2020.

4. Galloway SE, Paul P, MacCannell DR, et al. Emergence of SARS-CoV-2 B.1.1.7 Lineage - United States, December 29, 2020-January 12, 2021. MMWR Morb Mortal Wkly Rep. 2021;70(3):95–9.

5. Food and Drug Administration. Accelerated Emergency Use Authorization Summary: Curative SARS-CoV-2 Assay. Table 11. Available at: https://www.fda.gov/media/137089/download Accessed March 24, 2021.

6. Kojima N, Turner F, Slepnev V, et al. Self-Collected Oral Fluid and Nasal Swab Specimens Demonstrate Comparable Sensitivity to Clinician-Collected Nasopharyngeal Swab Specimens for the Detection of SARS-CoV-2. Clin Infect Dis. 2020.

7. Macias Gil R, Marcelin JR, Zuniga-Blanco B, Marquez C, Mathew T, Piggott DA. COVID-19 Pandemic: Disparate Health Impact on the Hispanic/Latinx Population in the United States. J Infect Dis. 2020.

8. Hellewell J, Abbott S, Gimma A, et al. Feasibility of controlling COVID-19 outbreaks by isolation of cases and contacts. Lancet Glob Health. 2020;8(4):e488–e96.

9. Allan-Blitz L-T, Klausner JD. Response to: Seroprevalence of SARS-CoV-2 following the largest initial epidemic wave in the United States: Findings from New York City, May 13-July 21, 2020. The Journal of Infectious Diseases. 2021.

10. Cerami C, Rapp T, Lin FC, et al. High household transmission of SARS-CoV-2 in the United States: living density, viral load, and disproportionate impact on communities of color. medRxiv. 2021.

11. Burr JA, Mutchler JE, Gerst K. Patterns of residential crowding among Hispanics in later life: immigration, assimilation, and housing market factors. J Gerontol B Psychol Sci Soc Sci. 2010;65(6):772–82.

12. Ogedegbe G, Ravenell J, Adhikari S, et al. Assessment of Racial/Ethnic Disparities in Hospitalization and Mortality in Patients With COVID-19 in New York City. JAMA Netw Open. 2020;3(12):e2026881.

13. Anand S, Montez-Rath M, Han J, et al. Prevalence of SARS-CoV-2 antibodies in a large nationwide sample of patients on dialysis in the USA: a cross-sectional study. Lancet. 2020.

14. Chen YH GM, Riley A, Balmes J, Duchowny Kate, Harrison R, Matthay E, Bibbins-Domingo K. Excess mortality associated with the COVID-19 pandemic among Californians 18–65 years of age, by occupational sector and occupation: March through October 2020. medRxiv. Available at: https://www.medrxiv.org/content/10.1101/2021.01.21.21250266v1.full.pdf. 2021.

15. Kaiser Family Foundation. State reports of long-term care facility cases and deaths related to COVID-19 (as of October 16, 2020). Accessed October 17, 2020. Available at: https://www.kff.org/health-costs/issue-brief/state-data-and-policy-actions-to-address-coronavirus/#top.

